# Enhancing Glaucoma Detection through Supervised Pre-training with Intermediate Phenotypes: A Multi-Institutional Study

**DOI:** 10.1101/2025.04.22.25326210

**Authors:** Yiheng Li, Francisco Carrillo-Perez, Adi Mohammed Al Owaifeer, Mohammed Alawad, Olivier Gevaert

## Abstract

Glaucoma is a leading cause of irreversible blindness worldwide, with early diagnosis often hindered by subtle symptomatology and the lack of comprehensive screening programs. In this study, we introduce a robust deep learning framework that leverages supervised pre-training with clinically relevant intermediate indicators, most notably the vertical cup-to-disc ratio (VCDR), to enhance glaucoma detection from color fundus images. Utilizing the expansive AIROGS dataset for pre-training, our multi-task learning strategy simultaneously addresses categorical diagnostic classification and VCDR regression. We evaluated three architectures, ResNet-18, DINOv2, and RETFound across multiple international cohorts, including DRISHTI, G1020, ORIGA, PAPILA, REFUGE1, and ACRIMA. Compared to out-of-domain pre-training or self-supervised pre-training, supervised pre-training achieved the best average performances on all of ResNet-18 (average AUROC = 0.857), DINOv2 (average AUROC = 0.788), and RETFound (average AUROC = 0.839). The best model is the ResNet-18 pre-trained with AIROGS diagnostic features, achieving the highest AUROC of 0.930 on the G1020 dataset, and an average AUROC of 0.857 across all datasets. These results underscore the superiority of incorporating domain-specific clinical labels to guide feature extraction, thereby improving model performance, and generalizability for glaucoma detection. Our findings advocate for the integration of supervised pre-training strategies into glaucoma detection model development, with significant potential to improve model performance and ultimately improve patient outcomes with decreased undiagnosed rate.

## INTRODUCTION

Glaucoma represents a major global health burden, ranking as the second leading cause of blindness worldwide and the most common cause of irreversible vision loss, affecting more than 70 million people globally (Li et al. 2023). Despite substantial advancements in ophthalmic care, glaucoma remains largely undiagnosed, with less than half of cases identified in high-income countries, and over 90% undetected in low- and middle-income countries (LMICs) (Burton et al. 2021). The primary reason for this low diagnosis rate is the subtlety and asymptomatic nature of early-stage glaucoma, often resulting in detection only at advanced stages when significant visual impairment or blindness has occurred, particularly in LMICs (Burton et al. 2021). Given these challenges, early detection and timely treatment are critical to preventing irreversible vision loss, underscoring the urgent need for scalable, effective screening solutions applicable across diverse patient populations (Burton et al. 2021).

Artificial intelligence (AI) has been widely applied in the medical field, particularly across various imaging modalities such as CT, MRI, histology image, etc, to optimize patient outcomes through predictive modeling (Li et al. 2024; Pizurica et al. 2024; Xu et al. 2024; Noor et al. 2024). In response to the urgent demand for scalable glaucoma screening, deep learning models utilizing Color Fundus Images (CFIs) have shown considerable promise by enabling automated, high-throughput glaucoma detection (Ling et al. 2025). Nevertheless, these models frequently encounter significant challenges in generalizing across diverse datasets. Variability in imaging devices, patient demographics, and inconsistencies in clinical annotation standards contribute to substantial domain shifts, diminishing model performance when deployed in real-world clinical scenarios (Orlando et al. 2020; Liu et al. 2019; Li et al. 2023). Achieving robust generalization across heterogeneous clinical settings thus remains a substantial hurdle for current AI approaches.

In order to deal with the generalizability issue, researchers found out that pre-training on larger and heterogeneous datasets is helpful. Among the pre-training methods, Self-Supervised Learning (SSL) methods have recently gained popularity in medical imaging and have been considered an important technique in building generalist medical AI due to their reduced reliance on extensive labeled datasets (Moor et al. 2023; C. Zhang, Zheng, and Gu 2023). However, evidence from recent studies indicates that SSL approaches sometimes rely on spurious image features or non-clinical artifacts rather than clinically relevant markers, or do not show sufficient morphological awareness for medical images, thus compromising their reliability in real-world clinical practice (Kauffmann et al. 2025; Yang Wena, Leiting Chena,b, Yu Dengc, Chuan Zhoua 2021). Such limitations necessitate alternative strategies that can better ensure the clinical relevance and robustness of model-derived insights. At the same time, supervised pre-training, particularly when integrated with clinical domain expertise, represents a compelling alternative capable of achieving clinically meaningful feature extraction and with improved robustness (Bear Don’t Walk et al. 2021; Schäfer et al. 2024). By leveraging supervised pre-training on clinically relevant intermediate indicators, such as the vertical cup-to-disc ratio (VCDR), a key anatomical marker strongly associated with glaucoma progression, models can better align feature learning with recognized clinical insights, thereby enhancing their predictive accuracy, interpretability, and generalizability across diverse patient cohorts (Tan et al. 2024; Gao et al. 2024).

To address these critical challenges in glaucoma, we introduce a supervised pre-training framework explicitly designed to enhance the robustness and clinical relevance of automated glaucoma detection (Figure 1). By explicitly integrating clinical domain knowledge through supervised pre-training, utilizing the diagnostic features as well as VCDR as intermediate clinical training signals, our model effectively guides feature learning towards clinically significant patterns, consequently improving both interpretability and generalization performance. Therefore, we rigorously evaluated our supervised learning approach against leading alternative methodologies, including established SSL techniques and traditional single-domain supervised learning approaches. Comparative analysis clearly demonstrates the superior robustness, predictive accuracy, and clinical relevance of our supervised pre-training strategy, as evidenced by consistent performance across diverse datasets and patient populations.

**Figure 1.**
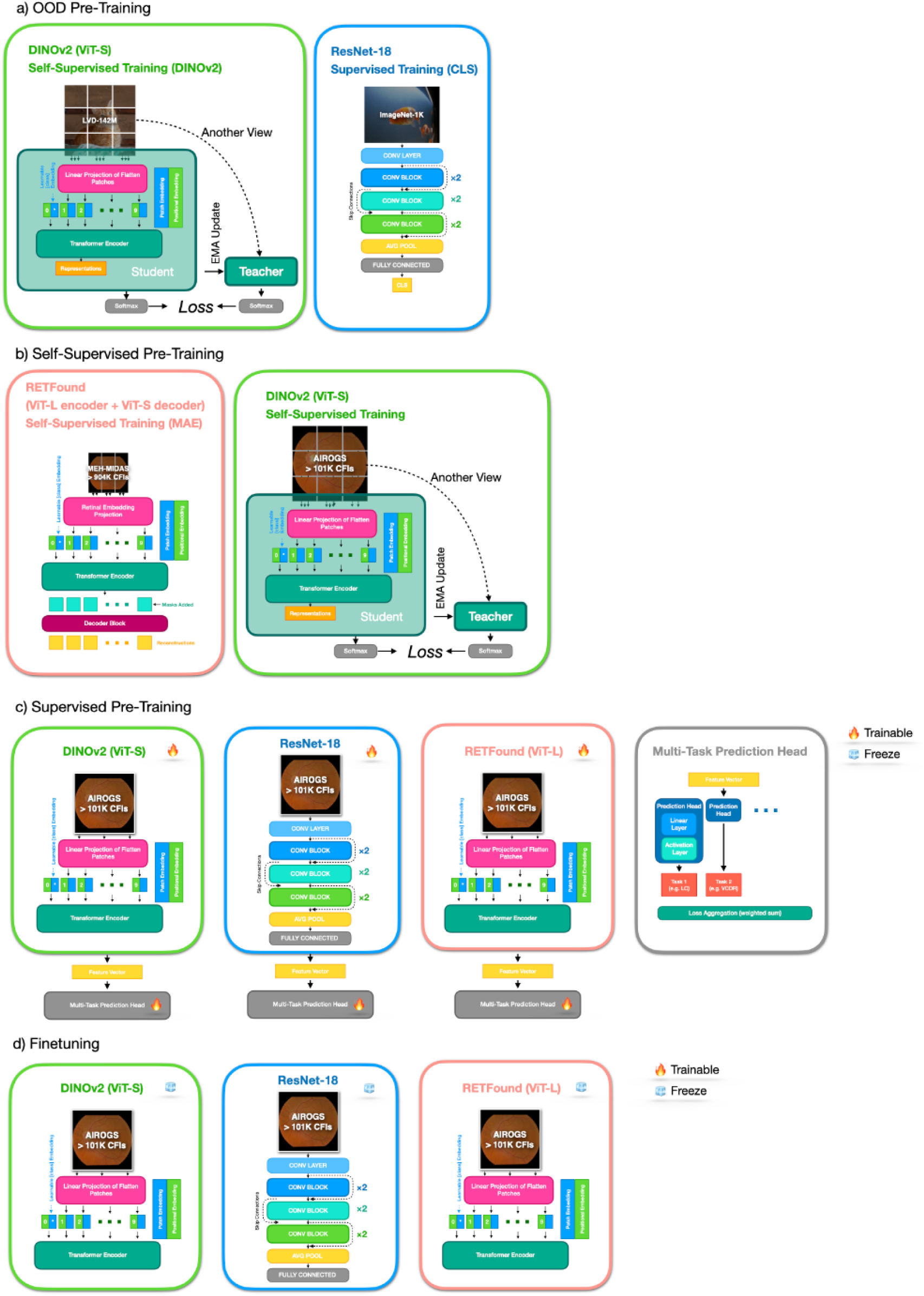
Overview of the pre-training and fine-tuning strategies of ResNet-18, DINOv2, and the RETFound models. a) out-of-domain (OOD) pre-training strategy; b) self-supervised learning (SSL) pre-training strategy; c) supervised pre-training strategy; d) Finetuning strategy.

The key contributions of our study are multifold: firstly, we demonstrate the effectiveness of a generalizable glaucoma classification system exhibiting consistently high accuracy and robustness across multiple diverse cohorts. Secondly, we provide strong empirical support for the superiority of supervised pre-training strategies over SSL and traditional methods. Thirdly, we innovatively utilize VCDR as an explicit supervised training signal, significantly enhancing clinical relevance and model interpretability. Lastly, our clinical impact evaluation quantitatively highlights the potential of our approach to substantially reduce undiagnosed glaucoma, establishing a compelling case for its implementation in clinical settings. Collectively, these contributions form a cohesive narrative that addresses the critical clinical need for improved glaucoma detection and validates our approach’s methodological innovation, robust validation, and practical significance.

## RESULTS

### Model Performance Across Cohorts

In our study, we employed three distinct model architectures for glaucoma classification: ResNet-18 (He et al. 2015) a residual CNN pre-trained on ImageNet; DINOv2 (Oquab et al. 2023) a self-supervised vision transformer trained on 142 million natural images; and RETFound (Y. Zhou et al. 2023) a masked autoencoder-based Vision Transformer pre-trained on 1.6 million retinal images and Optical Coherence Tomography (OCTs). To enhance model performance on glaucoma-related classification tasks, we first conducted supervised pre-training using the AIROGS dataset (de Vente et al. 2024) (Table 1), a large cohort comprising 112,732 CFIs from 60,071 subjects across 500 clinical sites, with diverse ethnic representations. AIROGS images were labeled by expert ophthalmologists and optometrists, whose assessments were validated through the European Optic Disc Assessment Trial (EODAT). The images were labeled with diagnostic features that are indicative of glaucoma as well as “referrable glaucoma” (Table S1). Fine-tuning and evaluation were performed through leave-one-dataset-out cross-validation across six distinct datasets, including five cohorts from the Standardized Multi-Channel Dataset for Glaucoma (SMDG-19) (Kiefer et al. 2023) and the standalone ACRIMA dataset (Diaz-Pinto et al. 2019) (Table 1). The SMDG-19 datasets were standardized to include full-fundus images, part of the images with segmentation data (i.e. optic disc, optic cup, and blood vessels), and demographic information (i.e. sex and age). The five cohorts (DRISHTI, G1020, ORIGA, PAPILA, REFUGE1) in the SMDG-19 dataset we chose have the clinically confirmed labeling of the disease of glaucoma. For data pre-processing, we tested whether standardizing the Field-of-View (FOV) of the fundus image would improve the result, but no consistent improvements were found, so we didn’t include it in the pipeline (Table S2). Besides the supervised pre-training, we also tested other pre-training strategies including Out-of-Domain (OOD) pre-training on natural images and self-supervised pre-training on CFIs (Figure 1), among the tested strategies the best-performing model was ResNet-18 (Table 2, Table 3), which, after supervised pre-training on AIROGS diagnostic features, achieved the highest AUROC of 0.930 on the G1020 dataset, a dataset part of SMDG-19, and an average AUROC of 0.857 across all evaluation datasets. The lowest performance was observed on the ACRIMA dataset (Table 2), where the model achieved an AUROC of 0.780, likely due to the dataset’s distinct visual characteristics, including cropped optic cup regions. Despite variations in dataset characteristics, the model demonstrated overall robustness and stability in glaucoma classification.

**Table 1.**
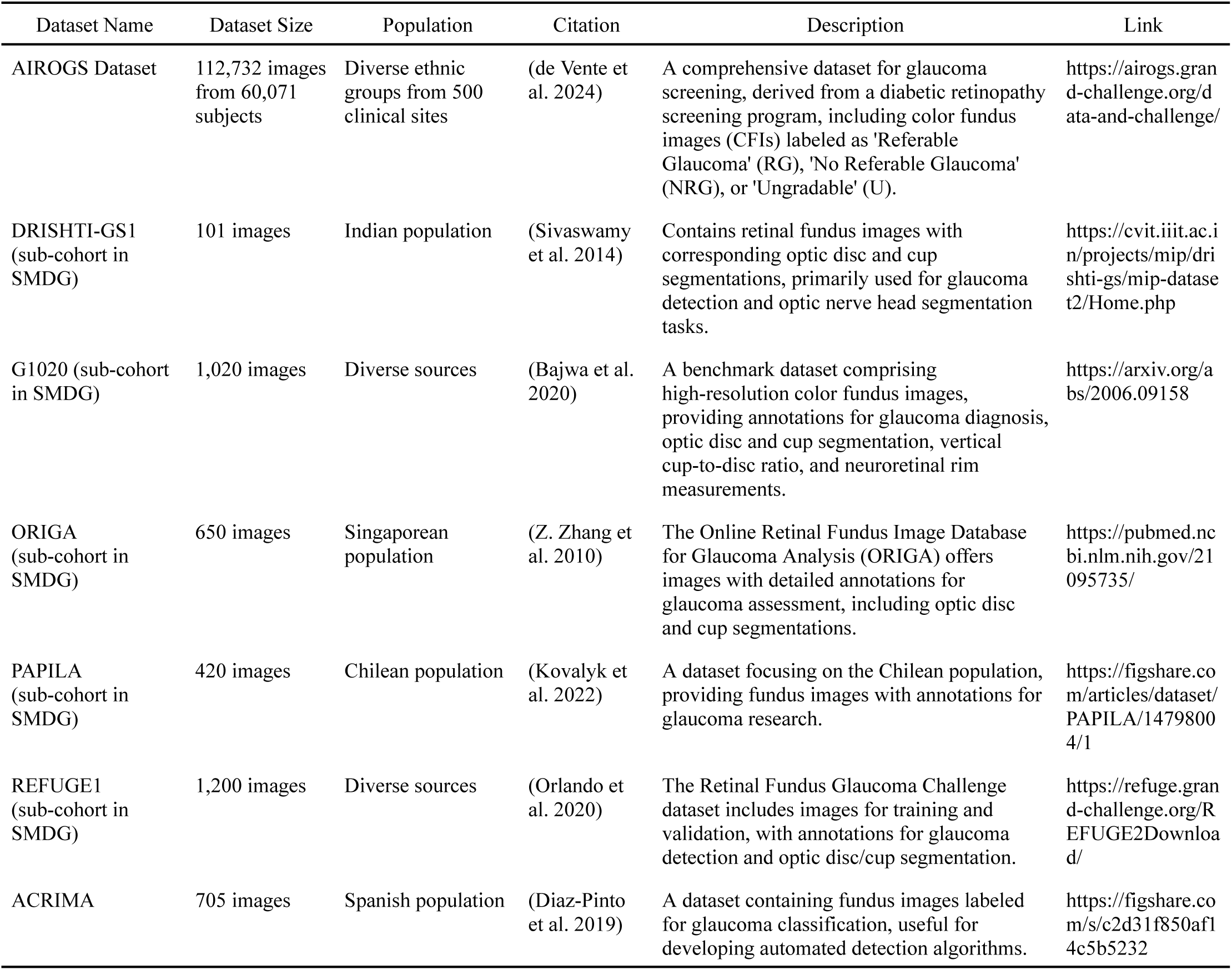
Overview of Datasets Used for Pre-training, Fine-Tuning, and Evaluation of the Glaucoma Classification.

**Table 2.** Performance of glaucoma classification using ResNet-18, pre-trained in a supervised manner on the AIROGS dataset and diagnostic features. The model is evaluated on 5 cohorts of the SMDG-19 (DRISHTI, G1020, ORIGA, PAPILA and REFUGE1) dataset and the ACRIMA dataset using a leave-one-dataset-out approach.

| Datasets | Accuracy | Precision | Recall | F1 Score | **AUROC** |
| --- | --- | --- | --- | --- | --- |
| DRISHTI | 0.821 | 0.724 | 0.600 | 0.656 | **0.882** |
| G1020 | 0.875 | 0.812 | 0.756 | 0.783 | **0.930** |
| ORIGA | 0.846 | 0.805 | 0.649 | 0.719 | **0.893** |
| PAPILA | 0.812 | 0.780 | 0.545 | 0.641 | **0.855** |
| REFUGE1 | 0.760 | 0.729 | 0.502 | 0.595 | **0.799** |
| ACRIMA | 0.732 | 0.442 | 0.543 | 0.487 | **0.780** |
| Average | 0.808 | 0.715 | 0.599 | 0.647 | **0.857** |

**Table 3.** Comparison of ResNet-18, DINOv2 and RETFound models with different pre-training methods. The table reports the AUROC for glaucoma classification. OOD is not relevant for RETFound, which is a private foundation model pre-trained using SSL on color fundus imaging. SSL is not compatible with ResNet-18.

| Datasets | OOD pre-training<br>on natural images |  |  | SSL pre-training<br>on color fundus<br>images |  |  | Supervised<br>pre-trained with<br>VCDR |  |  | Supervised<br>pre-trained with<br>AIROGS<br>diagnostic features |  |  | Supervised<br>pre-trained with<br>VCDR and<br>AIROGS<br>diagnostic features |  |  |
| --- | --- | --- | --- | --- | --- | --- | --- | --- | --- | --- | --- | --- | --- | --- | --- |
|  | ResNet-18 | DINOv2 | RETFound | ResNet-18 | DINOv2 | RETFound | ResNet-18 | DINOv2 | RETFound | ResNet-18 | DINOv2 | RETFound | ResNet-18 | DINOv2 | RETFound |
| DRISHTI | 0.818 | 0.807 | - | - | 0.754 | 0.828 | 0.864 | 0.815 | 0.847 | 0.882 | 0.793 | 0.807 | 0.869 | 0.809 | 0.849 |
| G1020 | 0.897 | 0.827 | - | - | 0.760 | 0.836 | 0.918 | 0.854 | 0.885 | 0.930 | 0.848 | 0.807 | 0.928 | 0.861 | 0.891 |
| ORIGA | 0.860 | 0.778 | - | - | 0.688 | 0.842 | 0.880 | 0.800 | 0.874 | 0.893 | 0.780 | 0.796 | 0.895 | 0.791 | 0.879 |
| PAPILA | 0.832 | 0.767 | - | - | 0.696 | 0.801 | 0.857 | 0.786 | 0.841 | 0.855 | 0.764 | 0.790 | 0.865 | 0.788 | 0.854 |
| REFUGE1 | 0.743 | 0.739 | - | - | 0.658 | 0.764 | 0.817 | 0.704 | 0.787 | 0.799 | 0.716 | 0.712 | 0.789 | 0.704 | 0.782 |
| ACRIMA | 0.702 | 0.693 | - | - | 0.687 | 0.567 | 0.772 | 0.754 | 0.766 | 0.780 | 0.748 | 0.805 | 0.774 | 0.774 | 0.780 |
| *Average* | **0.809** | 0.769 | - | - | 0.707 | 0.773 | **0.851** | 0.786 | 0.833 | **0.857** | 0.775 | 0.786 | **0.853** | 0.788 | 0.839 |

### Effect of Supervised Pre-training

We compared three pre-training strategies for glaucoma classification in color fundus images: (i) OOD pre-training on natural images, (ii) self-supervised learning (SSL) on CFIs, and (iii) supervised pre-training on color fundus images using VCDR and AIROGS diagnostic features. In the OOD setting, we used publicly available ResNet-18 weights, originally trained in a supervised fashion on ImageNet, and DINOv2, originally trained in a self-supervised manner on large-scale natural image datasets. Under these respective OOD initializations, ResNet-18 achieved an AUROC of 0.809, whereas DINOv2 reached 0.769 (Table 3). For SSL on CFIs, we applied the DINOv2 SSL framework to the AIROGS dataset in an unlabeled manner, obtaining an AUROC of 0.707. Similarly, RETFound was SSL trained on unlabeled MEH-MIDAS and EyePACS dataset, yielding an AUROC of 0.773 on the evaluation datasets. In supervised pre-training on the AIROGS dataset, we evaluated different combinations of intermediate labels - VCDR, AIROGS diagnostic features, or VCDR combining with AIROGS diagnostic features (Table S1). In supervised pre-training settings, the best performance of the ResNet-18 model is obtained when training with the AIROGS diagnostic features with averaged AUROC of 0.857 (Table 3). The best performing DINOv2 (average AUROC = 0.788) and RETFound (average AUROC = 0.839) in the setting of supervised pre-training were both achieved by training with VCDR and AIROGS diagnostic features combined. Across all architectures, supervised pre-training on color fundus images consistently delivered the highest performance (Table 3). These findings underscore the superiority of fundus-specific supervised pre-training over purely SSL-based or OOD approaches.

### Pre-training with Diagnostic Labels as well as VCDR

We leveraged intermediate labels consisting of categorical diagnostic labels (e.g. laminar dots, abnormal neuroretinal rim, and referable glaucoma, etc. Table S1) from expert assessments that are available in the AIROGS dataset, as well as calculated numerical vertical cup-to-disc ratio (VCDR) values. The VCDR was derived by first segmenting the optic disc and optic cup from the color fundus image using an automated deep learning model, SegFormer, which performed with 0.929 on optic discs and 0.804 on optic cups on the evaluation datasets that has the ground truth segmentation (Table S3). Subsequently, VCDR was calculated from these segmentations using a dedicated algorithm, as detailed in the supplementary material (Algorithm 2). Using a multi-task supervised pre-training framework, we set up the model to predict multiple classification tasks on the diagnostic labels of the AIROGS dataset, or solely predict regression of the VCDR, or jointly perform classification of the diagnostic labels and regression of VCDR. For the choice of predicting the VCDR as a classification or regression task, we tried predicting VCDR as a classification task by splitting the VCDR into two categories, but it yielded worse results compared to regression the VCDR, so we continued with treating the VCDR as a regression label (Table S4). After training the models to predict the intermediate tasks of diagnostic labels or VCDR, we obtained the model’s performance on the validation split of the AIROGS dataset (Table S5, S6 a–f). Then, we fine-tuned the models to predict the glaucoma detection task. The glaucoma detection performances were gathered on all evaluation datasets by leaving one dataset out for evaluation and trained on the rest of the datasets. ResNet-18 attained the highest average AUROC (0.857) and peak AUROC (0.930) when trained solely on the categorical AIROGS diagnostic features. In contrast, incorporating VCDR labels was pivotal for DINOv2, compared to training on AIROGS features, training with VCDR increased the average AUROC from 0.775 to 0.786 and peak AUROC from 0.848 to 0.854, both on the G1020 dataset. Combining both the VCDR and the AIROGS features gives the best performance, with the average AUROC being 0.788. Similarly, RETFound achieved higher average AUROC when training with VCDR (0.833) compared to AIROGS features (0.786), and the highest average AUROC is achieved by combining both the AIROGS features and the VCDR (0.839) (Table 3). These findings illustrate the consistent benefit of integrating both categorical and numerical intermediate labels, particularly VCDR, in elevating performance and enhancing model robustness across multiple architectures.

## DISCUSSION

This study presents a deep learning model designed explicitly for glaucoma detection, evaluated across diverse international cohorts. Our findings demonstrate that supervised pre-training strategies significantly enhance the performance and generalizability of deep learning models compared to purely SSL methods or models pre-trained on out-of-domain datasets. A key insight from our work is the substantial advantage provided by supervised pre-training, even in scenarios lacking extensive expert labeling, through the use of computationally derived clinical indicators, such as the VCDR. While SSL methods have garnered significant interest for foundation model development due to their scalability and reduced labeling requirements, our comparative analyses clearly demonstrate that supervised pre-training strategies remain highly competitive or superior in medical imaging contexts, particularly for glaucoma detection. Our results align with recent studies that similarly underscore the clinical utility and effectiveness of supervised approaches for specific medical problems, highlighting the enduring importance of integrating domain-specific clinical knowledge into model training (Chen, Li, and Wan 2022).

Previous research has established VCDR as a critical anatomical marker associated with glaucoma progression, often incorporating VCDR directly as input features for glaucoma prediction models or as an independent predictor for glaucoma (Hemelings et al. 2023; Gao et al. 2024). Our study innovatively repurposed VCDR as an intermediate supervised training signal during pre-training, effectively guiding the deep learning model to learn clinically meaningful patterns relevant to glaucoma pathology. Our experiments validate this strategy, showing that incorporating VCDR significantly improved both predictive performance and model interpretability. Moreover, our findings underscore the feasibility of fully automating VCDR calculation through deep learning-based segmentation algorithms, thus enabling large-scale deployment without manual labeling burdens. This approach exemplifies a promising paradigm where outputs from one deep learning model (i.e. segmentation-based VCDR calculation) serve as valuable training inputs for subsequent predictive deep learning models, creating a beneficial feedback loop that enhances overall diagnostic accuracy and efficiency.

Our methodological framework aligns closely with recent advancements such as GONet and G-RISK, which similarly leverage anatomical features for glaucoma risk assessment (Hemelings et al. 2023; Gao et al. 2024). However, our study uniquely demonstrates the substantial benefits derived specifically from supervised pre-training strategies using intermediate clinical indicators like VCDR, thereby extending and refining the utility of these prior methods.

In conclusion, our study provides significant contributions by introducing a highly generalizable glaucoma classification system robustly validated across multiple diverse patient cohorts. We clearly establish the superiority of supervised pre-training methods, leveraging clinically relevant intermediate indicators like VCDR, over purely SSL or traditional OOD pre-training strategies. Additionally, our innovative use of VCDR as an explicit supervised training signal not only enhances predictive accuracy and model interpretability but also facilitates a fully automated pipeline suitable for large-scale clinical deployment. Collectively, these contributions address critical clinical needs in glaucoma detection, offering methodological innovations, robust validation strategies, and substantial potential for impactful clinical integration.

## CODE AVAILABILITY

The code as well as model weights will be provided when this work is officially published.

## DATA AVAILABILITY

The datasets used in this study are all publicly available. The AIROGS dataset can be accessed through https://airogs.grand-challenge.org/. The SMDG-19 dataset can be accessed through https://www.kaggle.com/datasets/deathtrooper/multichannel-glaucoma-benchmark-dataset. The ACRIMA dataset can be accessed through https://www.kaggle.com/datasets/toaharahmanratul/acrima-dataset.

## METHODS

### Data and Cohorts

For this study, the Rotterdam EyePACS AIROGS dataset served as our primary resource for pre-training. AIROGS comprises 112,732 color fundus images (CFIs) from 60,071 subjects across 500 clinical sites, featuring a diverse ethnic distribution. The images were collected during a diabetic retinopathy screening initiative, with expert ophthalmologists and optometrists, trained through the European Optic Disc Assessment Trial (EODAT), assigning each CFI to one of three categories: “Referable Glaucoma” (RG), “No Referable Glaucoma” (NRG), or “Ungradable” (U). Only RG and NRG images were used in this analysis. The AIROGS dataset was then split into a training set of 101,442 CFIs and a test set of 11,290 CFIs, ensuring patient-level independence. We further split the training set into a validation set of 19,848 images, and the remaining images are served as training images.

To evaluate the performance and generalizability of our models, we additionally employed the ACRIMA dataset and a subset of the Standardized Multi-Channel Dataset for Glaucoma (SMDG-19). ACRIMA, collected at FISABIO Oftalmología Médica in Valencia, Spain, contains 705 CFIs (396 glaucoma, 309 normal) obtained under ethical standards consistent with the Helsinki Declaration. Each image was annotated by glaucoma specialists, making the dataset suitable for both validation of diagnostic algorithms and medical education (Table 3). Meanwhile, SMDG-19 consolidates and standardizes images from 19 publicly available glaucoma datasets; for this study, we specifically focused on five clinically validated datasets (DRISHTI, G1020, PAPILA, REFUGE1, and ORIGA) that provide confirmed glaucoma labels (Table 3). Each image underwent standardized preprocessing, dynamic global foreground thresholding followed by uniform resizing to 512×512 pixels, to enable consistent binary glaucoma classification across all evaluation datasets.

### Model Architectures

In our study, we employed three distinct model architectures, ResNet-18, DINOv2, and RETFound, each leveraging unique pre-training strategies to enhance performance in ophthalmic image analysis. (Figure 1)

**ResNet-18** (He et al. 2015): This 18-layer convolutional neural network is renowned for its efficacy in image classification tasks. We utilized a version pre-trained on the ImageNet dataset, which comprises over a million images across 1,000 categories. This pre-training enables the model to learn rich feature representations applicable to a wide array of images.

**DINOv2** (Oquab et al. 2023): Developed by Meta AI Research, DINOv2 is a self-supervised Vision Transformer (ViT) model. It was trained on a curated dataset of 142 million images from diverse sources without using any labels or annotations, allowing it to learn high-performance visual features that can be directly employed with classifiers as simple as linear layers on various computer vision tasks.

**RETFound** (Zhou et al. 2023): This model is based on a masked autoencoder architecture featuring a ViT-large encoder and a ViT-small decoder. It was pre-trained on a substantial dataset comprising 904,170 CFIs and 736,442 optical coherence tomography (OCT) scans from diverse sources, primarily the Moorfields Diabetic Image Dataset (MEH-MIDAS). This extensive pre-training enables RETFound to learn rich visual representations pertinent to ophthalmic imaging.

### Pre-Training Strategy

In our study, we implemented multiple pre-training strategies tailored to optimize model performance for ophthalmic image analysis. These included OOD pre-training on natural images, SSL on domain-specific fundus images, and supervised pre-training incorporating diagnostic labels and VCDR values.

#### Out-of-Domain Pre-Training

OOD pre-training involved initializing models with weights obtained from large-scale natural image datasets, allowing them to learn fundamental visual representations before being adapted to ophthalmic tasks. ResNet-18 was originally trained using supervised learning (SL) on the ImageNet dataset, which consists of over one million images spanning 1,000 object categories. This pre-training enabled ResNet-18 to capture hierarchical spatial features relevant to a broad range of image classification tasks (He et al. 2015). Unlike ResNet-18, DINOv2 was pre-trained in a SSL manner on a curated collection of 142 million natural images. This approach followed a teacher-student self-distillation framework, where the model learned to generate consistent representations across augmented views of the same image without requiring explicit human-provided labels (Oquab et al. 2023). By leveraging SSL, DINOv2 developed rich, transferable feature representations applicable to various computer vision domains.

#### Self-Supervised Learning on Domain-Specific Images

To further adapt models to the ophthalmic imaging domain, additional self-supervised pre-training was conducted using large-scale color fundus image datasets. We applied SSL to the AIROGS dataset, following the same pre-training framework as its original implementation. Specifically, we used a teacher-student self-distillation strategy, wherein a momentum-based teacher network generated target embeddings that the student network learned to predict. This approach enabled DINOv2 to refine its representations specifically for fundus images, capturing anatomical structures relevant to glaucoma assessment without relying on manual annotations. Unlike DINOv2, RETFound was pre-trained on domain-specific images from the outset using a masked autoencoder (MAE) framework. This SSL pre-training utilized a dataset of 904,170 color fundus images (CFIs) and 736,442 optical coherence tomography (OCT) scans, primarily sourced from the Moorfields Diabetic Image Dataset (MEH-MIDAS), with additional images from Kaggle EyePACS and other sources. RETFound’s encoder, a ViT-large model, processed partially masked image patches and learned to reconstruct the missing regions, encouraging it to develop strong feature representations for retinal structures. The pre-training phase was done by the original authors of RETFound (Y. Zhou et al. 2023). It lasted 800 epochs with a batch size of 1,792 across eight NVIDIA A100 GPUs, ensuring robust feature extraction capabilities for ophthalmic imaging tasks.

#### Supervised Pre-Training with Diagnostic Labels and VCDR

In addition to self-supervised learning, we conducted supervised pre-training using structured diagnostic labels and automatically derived numerical features. Specifically, we leveraged diagnostic labels from the AIROGS dataset, along with VCDR values calculated using a SegFormer (Diaz-Pinto et al. 2019; Xie et al. 2021) segmentation algorithm and a VCDR calculation algorithm, inspired by S. Zhou’s algorithm (S. Zhou et al. 2021), as detailed in the supplementary material. This setup facilitated multi-task learning, where the models were trained to perform both classification (diagnostic label prediction) and regression (VCDR estimation). Each task was assigned a separate linear layer: the regression task used sigmoid activation and SmoothL1Loss, while the classification task used no activation and CrossEntropyLoss. By integrating both categorical and numerical glaucoma indicators, supervised pre-training enhanced the models’ ability to capture disease-related patterns in fundus images.

### Multi-task Learning Strategy

We implemented a multi-task learning strategy to simultaneously predict multiple glaucoma-related outcomes, integrating both regression and classification tasks to optimize shared model representations. The primary regression task involved predicting the vertical cup-to-disc ratio (VCDR), a key quantitative indicator of glaucoma severity, using SmoothL1Loss with a weighting factor of 10.0 to balance its contribution within the overall loss function. In parallel, the classification branch included binary classification of referable glaucoma (RG vs. NRG) as well as several clinically relevant indicators linked to structural and pathological changes in the optic nerve head. These indicators included neuroretinal rim abnormalities (ANRS and ANRI), superior and inferior retinal nerve fiber layer defects (RNFLDS and RNFLDI), baying of circumlinear vessels (BCLVS and BCLVI), neuroretinal rim visibility at the temporal region (NVT), disc hemorrhages (DH), laminar dots (LD), and large optic disc cupping (LC) (Table 4). Each classification task was trained using CrossEntropyLoss with a weight of 1.0. The total loss function was formulated as a weighted sum of all individual task losses, allowing the model to leverage shared representations and improve generalizability across multiple glaucoma-related features.

## SUPPLEMENTARY MATERIALS

**Table S1.**
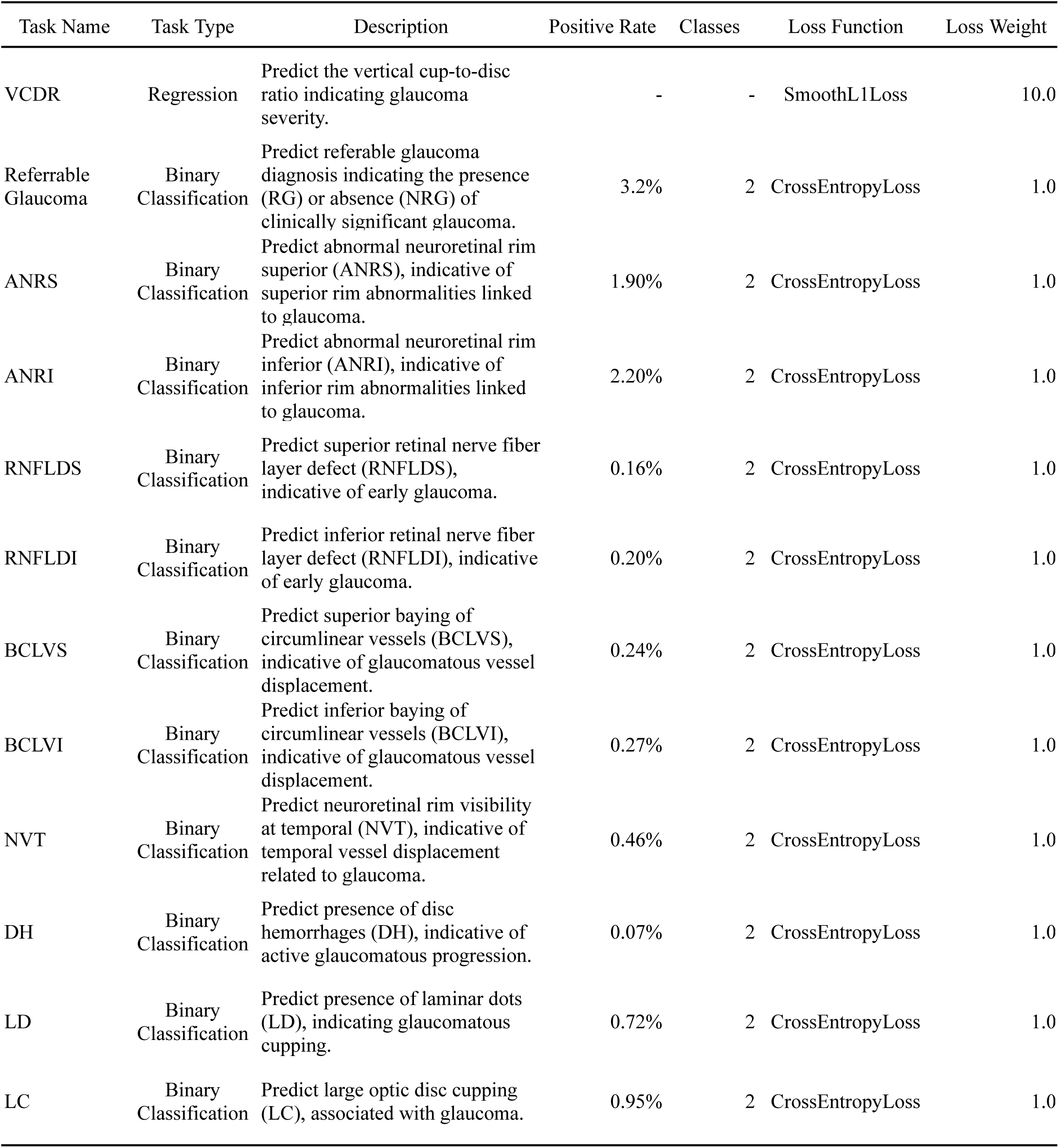
Overview of Diagnostic Labels in the AIROGS Dataset Used as Supervised Pre-training Labels for Glaucoma Prediction Models.

### Algorithm 1. VCDR Calculation from Segmented Images

**Input:**

- **cup_arr**: Binary 2D array, where pixels corresponding to the optic cup are marked as 1.
- **disc_arr**: Binary 2D array, where pixels corresponding to the optic disc are marked as 1.
- **eye**: Character (‘R’ or ‘L’) indicating the image’s laterality (right or left eye).

**Output:**

- **VCDR**: Ratio of the vertical cup diameter to the vertical disc diameter.
- **HCDR**: Ratio of the horizontal cup diameter to the horizontal disc diameter.
- **ISNT Metrics**: Neuroretinal rim thickness measurements in the inferior, superior, nasal, and temporal quadrants (ISNT_I, ISNT_S, ISNT_N, ISNT_T).
- **DDLS Score**: Disc Damage Likelihood Scale score derived from the minimum rim width over various rotated orientations.
- **Other Metrics**: Vertical and horizontal diameters for cup and disc, minimum rim width, and the angle at which this minimum occurs.

**Procedure:**

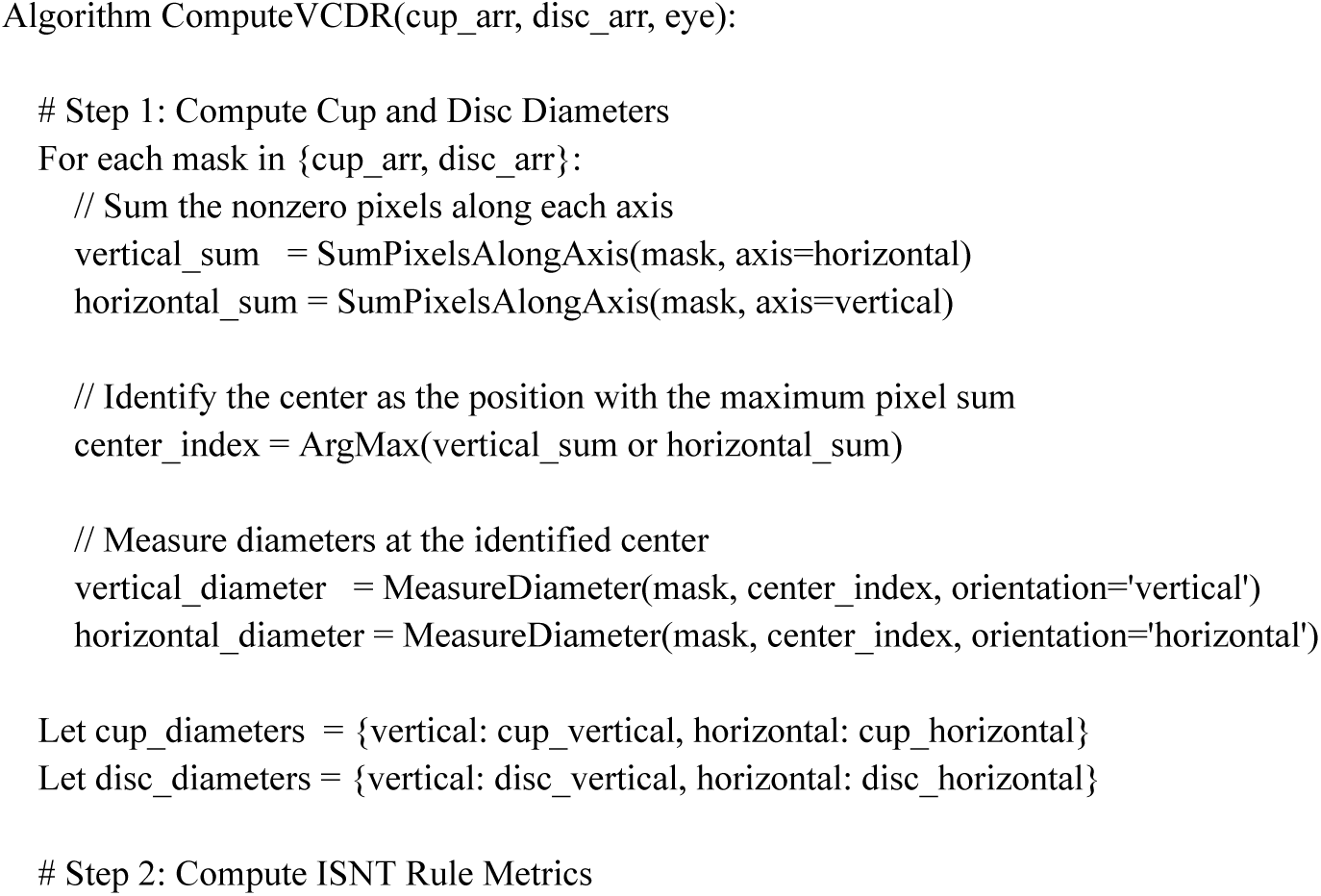
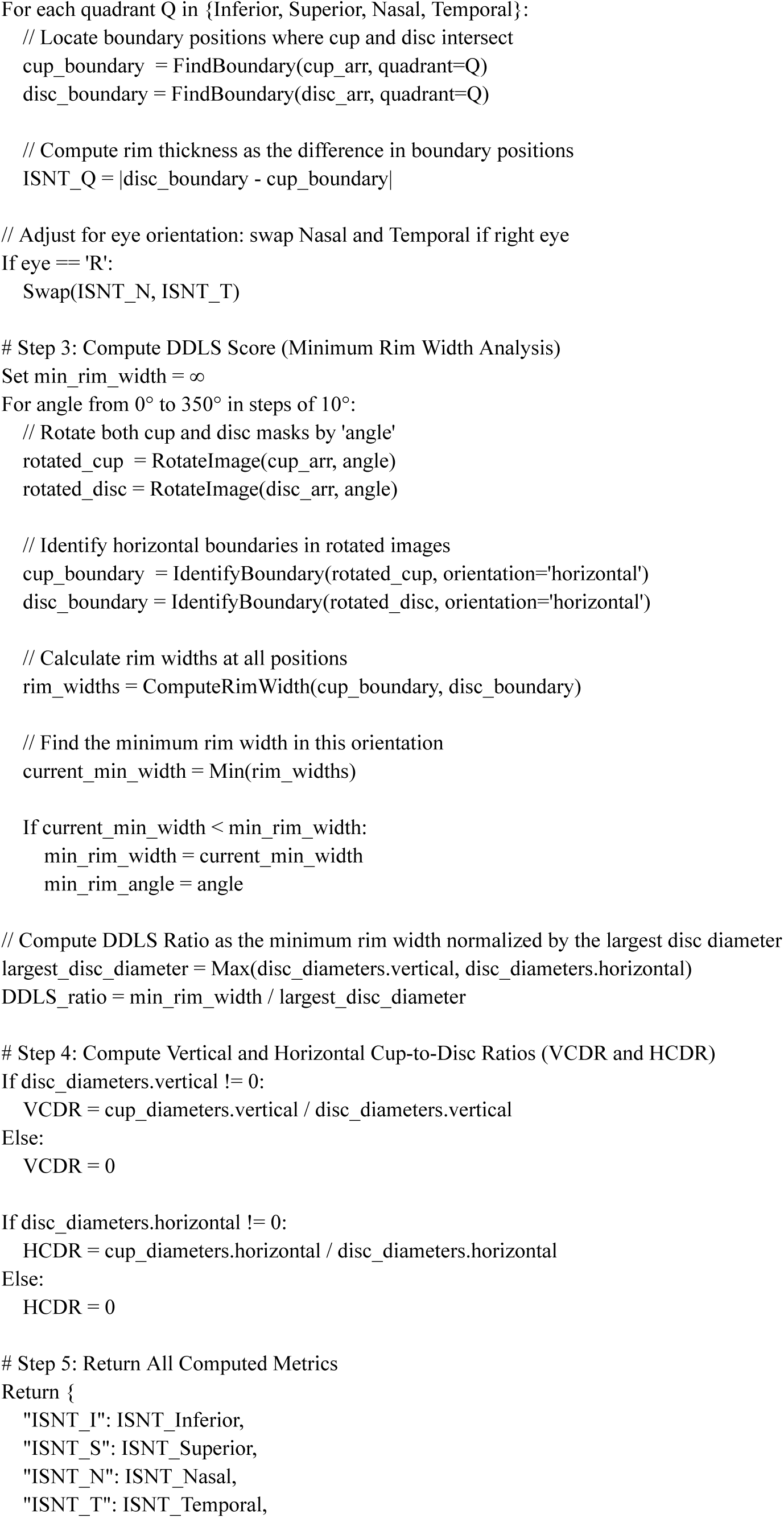
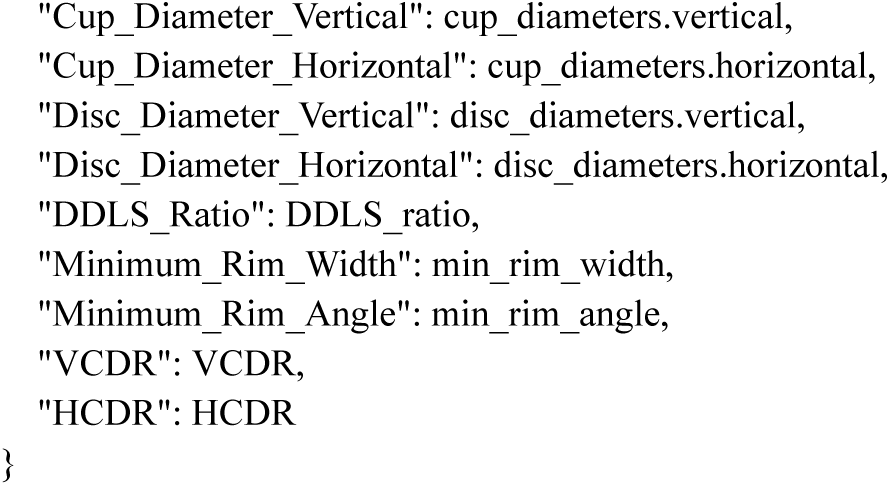

### Algorithm 2. Standardized Image Processing for Field of View (FOV) Adjustment

**Input:**

- **I**: Color image with dimensions H × W.
- **M**: Binary mask indicating the region of interest.
- **FOV_degrees**: Desired field of view in degrees.

**Output:**

- **Canvas**: A processed 512 × 512 image with the region of interest standardized.

**Procedure:**

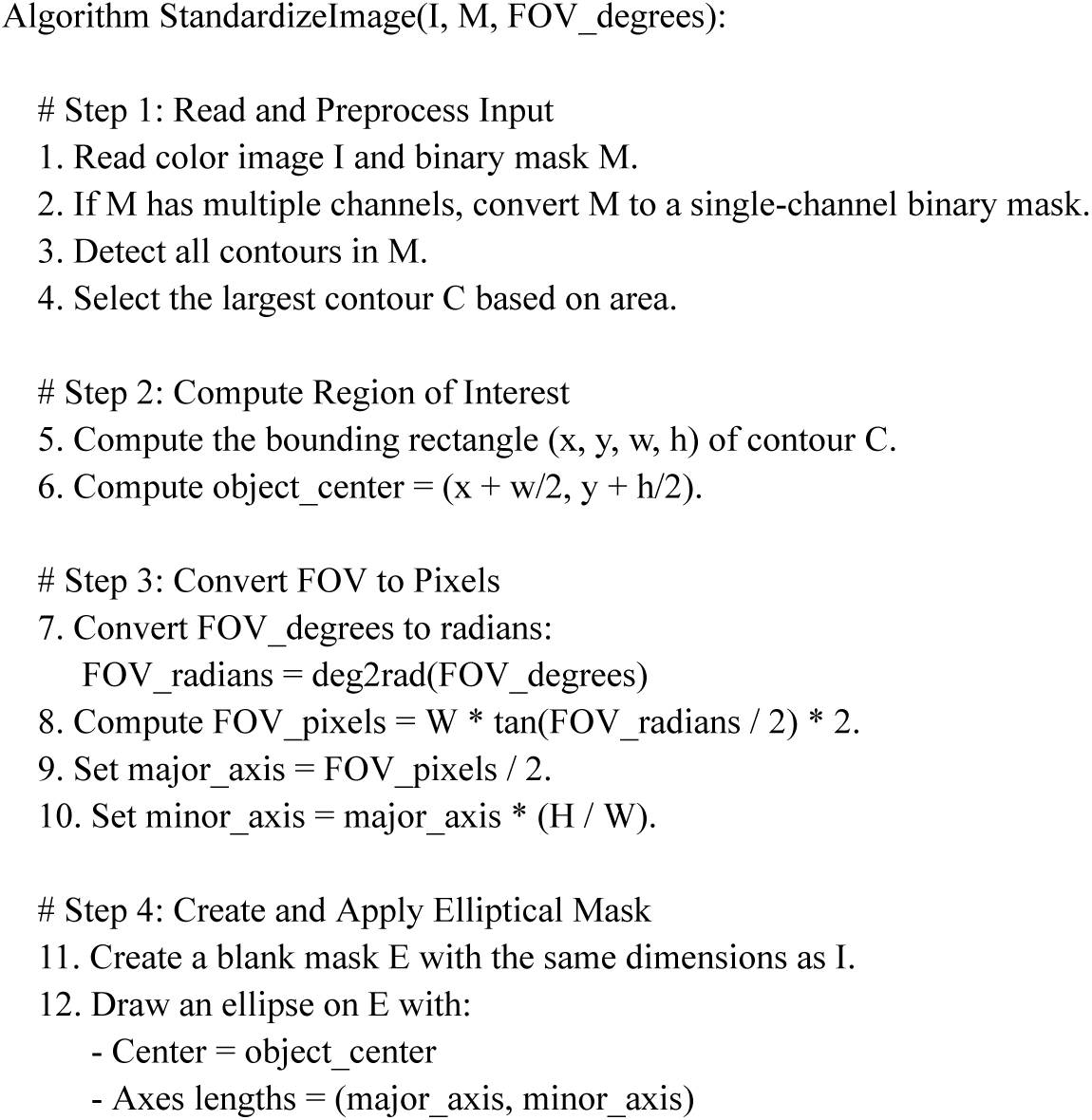
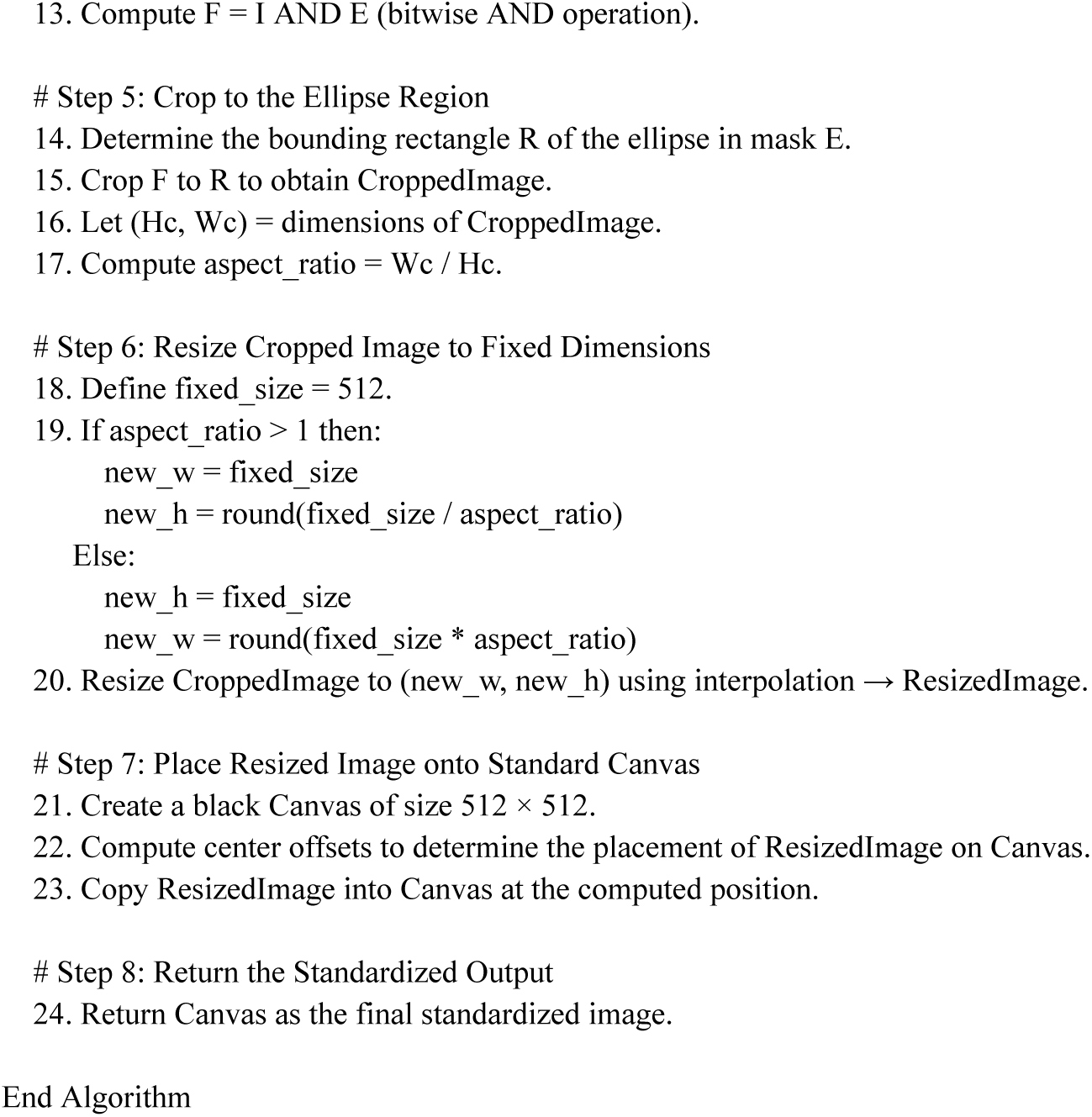

### Field-of-View Preprocessing

In this ablation study, we examined the impact of incorporating FOV preprocessing on glaucoma detection performance across three distinct deep learning models: ResNet-18, DINOv2, and RETFound. The effectiveness of FOV preprocessing was assessed by comparing the Area Under the Receiver Operating Characteristic Curve (AUROC) scores obtained with and without FOV preprocessing across six of our evaluation datasets (DRISHTI, G1020, ORIGA, PAPILA, REFUGE1, and ACRIMA) (Table S2).

**Table S2.** Model Classification Performance (AUROC) with or without FOV processed in the evaluation datasets.

| Datasets | ResNet-18 (ImageNet Pre-trained)<br>FOV preprocessing / control | DINOv2 (ImageNet Pre-trained)<br>FOV preprocessing / control | RETFound<br>FOV preprocessing / control |
| --- | --- | --- | --- |
| DRISHTI | 0.818 / **0.822** | **0.807** / 0.788 | **0.828** / 0.816 |
| G1020 | **0.897** / 0.863 | **0.827** / 0.807 | **0.836** / 0.815 |
| ORIGA | 0.860 / **0.862** | 0.778 / **0.780** | **0.842** / 0.831 |
| PAPILA | 0.832 / **0.839** | 0.767 / **0.779** | **0.801** / 0.786 |
| REFUGE1 | 0.743 / **0.764** | **0.739** / 0.693 | **0.764** / 0.740 |
| ACRIMA | **0.702** / 0.689 | 0.693 / **0.702** | 0.567 / **0.655** |

Across the datasets, the impact of FOV preprocessing varied depending on both the dataset and the specific model architecture employed. For ResNet-18, FOV preprocessing significantly enhanced performance on the G1020 and ACRIMA datasets but slightly reduced performance on ORIGA, PAPILA, and REFUGE1. The DINOv2 model benefited from FOV preprocessing, showing notable improvements on DRISHTI, G1020, and REFUGE1 datasets. RETFound exhibited improved performance with FOV preprocessing on most datasets except for ACRIMA, where a substantial drop in AUROC was observed. Overall, FOV preprocessing improved performance in some dataset–model combinations, it did not consistently yield benefits across all settings, so we ultimately chose not to include it in the final pipeline.

### Segmentation Performance of Optic Disc and Cup Using Segformer

We employed the Segformer model to segment the optic disc and cup in color fundus images as a necessary step toward computing the VCDR. The model was trained on the AIROGS dataset and evaluated on multiple subsets of the SMDG dataset, each containing ground truth segmentation masks. Performance was quantified using the Dice similarity coefficient for both the optic disc and cup (Table S3).

**Table S3.** Dice scores (mean ± standard deviation) of Segformer-based segmentation for optic disc and cup across SMDG subsets.

| Dataset | Dice Score for Optic Disc | Dice Score for Optic Cup |
| --- | --- | --- |
| REFUGE1 | 0.958 ± 0.016 | 0.895 ± 0.049 |
| G1020 | 0.919 ± 0.079 | 0.814 ± 0.122 |
| ORIGA | 0.938 ± 0.050 | 0.798 ± 0.111 |
| DRISHTI | 0.960 ± 0.027 | 0.653 ± 0.112 |
| PAPILA | 0.877 ± 0.234 | 0.677 ± 0.264 |
| CRFO | 0.947 ± 0.069 | 0.785 ± 0.187 |
| **Overall** | **0.929 ± 0.112** | **0.804 ± 0.162** |

Segformer achieved high segmentation performance for the optic disc across all datasets, with Dice scores exceeding 0.91 in most cases. The best performance was observed on DRISHTI (0.960) and REFUGE1 (0.958). In contrast, cup segmentation showed greater variability across datasets. While REFUGE1 (0.895) and G1020 (0.814) yielded strong results, performance was lower on DRISHTI (0.653) and PAPILA (0.677). These findings demonstrate the robustness of Segformer for optic disc segmentation and highlight opportunities for improvement in cup segmentation, especially in heterogeneous datasets.

### VCDR as a Regression Task vs. a Classification Task

**Table S4.** Model Classification Performance (AUROC) in the evaluation datasets after pre-training by predicting VCDR as a regression task or classification task in the AIROGS dataset datasets.

| Datasets | ResNet-18 (AIROGS features + VCDR Pre-trained)<br>regression / classification | DINOv2 (AIROGS features + VCDR Pre-trained)<br>regression / classification | RETFound (AIROGS features + VCDR Pre-trained)<br>regression / classification |
| --- | --- | --- | --- |
| DRISHTI | **0.869** / 0.818 | **0.815** / 0.799 | **0.849** / 0.791 |
| G1020 | **0.928** / 0.835 | **0.854** / 0.830 | **0.891** / 0.781 |
| ORIGA | **0.895** / 0.851 | 0.800 / **0.811** | **0.879** / 0.779 |
| PAPILA | **0.865** / 0.795 | 0.786 / **0.794** | **0.854** / 0.763 |
| REFUGE1 | **0.789** / 0.765 | **0.704** / 0.691 | **0.782** / 0.691 |
| ACRIMA | **0.774** / 0.641 | **0.754** / 0.563 | **0.780** / 0.277 |

To assess the impact of formulating the VCDR as a regression versus classification task, we conducted an ablation study across six external datasets using three different backbone models (ResNet-18, DINOv2, and RETFound). All models were pre-trained on the AIROGS dataset using a multi-task learning framework that included glaucoma-related classification tasks. For each backbone, we compared two variants: one with VCDR modeled as a regression task (using SmoothL1Loss) and another with VCDR modeled as a classification task with a threshold of 0.7 (using CrossEntropyLoss after discretizing VCDR into two groups, low and high).

Across all models and datasets, modeling VCDR as a regression task generally yielded superior performance in downstream glaucoma detection compared to its classification counterpart. The advantage of the regression formulation was particularly evident in datasets with clinically confirmed VCDR annotations, such as G1020 and ORIGA. For instance, ResNet-18, when pre-trained with VCDR as a regression task, achieved markedly higher performance on G1020 (0.928 vs. 0.835 for classification) and ORIGA (0.895 vs. 0.851). Similarly, DINOv2 demonstrated notable improvements from regression-based pre-training, especially on G1020 (0.854 vs. 0.830) and DRISHTI (0.815 vs. 0.799), although slight performance decreases were observed on ORIGA and PAPILA. RETFound, despite being the strongest model overall, also benefited from the regression formulation, showing consistent gains on DRISHTI (0.849 vs. 0.791), G1020 (0.891 vs. 0.781), and ORIGA (0.879 vs. 0.779). In contrast, the performance differences were less pronounced in datasets with noisier or less reliable VCDR annotations, such as REFUGE1 and ACRIMA, underscoring the importance of high-quality continuous VCDR labels for effective regression-based supervision (Table S4).

### The Model’s Performance on the Supervised Learning Intermediate Tasks

To assess the effectiveness of the supervised pre-training, after training on the AIROGS dataset, we also tested the model’s performance on predicting the supervised pre-trained task. This evaluation is done on 19,848 images of the dataset (~20%), randomly split as an evaluation set (Table S5, S6).

**Table S5.** VCDR regression results for models trained with VCDR on the AIROGS dataset.

| Models | MSE | R2 |
| --- | --- | --- |
| ResNet-18 trained on VCDR | 0.00375 | 0.543 |
| DINOv2 trained on VCDR | 0.00271 | 0.670 |
| RETFound trained on VCDR | 0.00287 | 0.650 |
| ResNet-18 trained on AIROGS features and VCDR | 0.00597 | 0.215 |
| DINOv2 trained on AIROGS features and VCDR | 0.00336 | 0.557 |
| RETFound trained on AIROGS features and VCDR | 0.00327 | 0.570 |

**Table S6a.** Diagnostic labels classification results for models trained with AIROGS diagnostic features for ResNet-18 model trained on AIROGS features.

| Diagnostic Features | Accuracy | Precision | Recall | F1 | AUROC | Confusion Matrix (tp/fn/fp/tn) |
| --- | --- | --- | --- | --- | --- | --- |
| ANRI | 0.986 | 0.216 | 0.313 | 0.256 | 0.930 | 46/101/167/19534 |
| ANRS | 0.989 | 0.256 | 0.419 | 0.318 | 0.937 | 52/72/151/19573 |
| BCLVI | 0.998 | 0.059 | 0.053 | 0.056 | 0.874 | 1/18/16/19813 |
| BCLVS | 0.999 | 0.000 | 0.000 | 0.000 | 0.735 | 0/23/3/19822 |
| DH | 0.999 | 0.000 | 0.000 | 0.000 | 0.772 | 0/9/1/19838 |
| Referrable Glaucoma | 0.982 | 0.295 | 0.452 | 0.357 | 0.929 | 98/119/234/19397 |
| LC | 0.993 | 0.200 | 0.205 | 0.202 | 0.933 | 17/66/68/19697 |
| LD | 0.996 | 0.143 | 0.115 | 0.128 | 0.835 | 6/46/36/19760 |
| NVT | 0.998 | 0.118 | 0.065 | 0.083 | 0.920 | 2/29/15/19802 |
| RNFLDI | 0.999 | 0.091 | 0.067 | 0.077 | 0.812 | 1/14/10/19823 |
| RNFLDS | 0.999 | 0.091 | 0.067 | 0.077 | 0.799 | 1/14/10/19823 |

**Table S6b.** Diagnostic labels classification results for models trained with AIROGS diagnostic features for ResNet-18 model trained on AIROGS features and VCDR.

| Diagnostic Features | Accuracy | Precision | Recall | F1 | AUROC | Confusion Matrix (tp/fn/fp/tn) |
| --- | --- | --- | --- | --- | --- | --- |
| ANRI | 0.977 | 0.109 | 0.311 | 0.162 | 0.885 | 28/62/228/12380 |
| ANRS | 0.976 | 0.113 | 0.459 | 0.182 | 0.933 | 34/40/266/12358 |
| BCLVI | 0.997 | 0.053 | 0.067 | 0.059 | 0.881 | 1/14/18/12665 |
| BCLVS | 0.997 | 0.040 | 0.067 | 0.050 | 0.860 | 1/14/24/12659 |
| DH | 0.999 | 0.000 | 0.000 | 0.000 | 0.689 | 0/8/3/12687 |
| Referrable Glaucoma | 0.951 | 0.113 | 0.538 | 0.186 | 0.902 | 71/61/560/12006 |
| LC | 0.986 | 0.082 | 0.222 | 0.120 | 0.914 | 12/42/134/12510 |
| LD | 0.992 | 0.040 | 0.094 | 0.056 | 0.804 | 3/29/72/12594 |
| NVT | 0.996 | 0.028 | 0.048 | 0.035 | 0.908 | 1/20/35/12642 |
| RNFLDI | 0.999 | 0.000 | 0.000 | 0.000 | 0.814 | 0/8/8/12682 |
| RNFLDS | 0.999 | 0.000 | 0.000 | 0.000 | 0.623 | 0/9/4/12685 |

**Table S6c.** Diagnostic labels classification results for models trained with AIROGS diagnostic features for the DINOv2 model trained on AIROGS features.

| Diagnostic Features | Accuracy | Precision | Recall | F1 | AUROC | Confusion Matrix (tp/fn/fp/tn) |
| --- | --- | --- | --- | --- | --- | --- |
| ANRI | 0.992 | 0.458 | 0.075 | 0.129 | 0.948 | 11/136/13/19688 |
| ANRS | 0.994 | 0.407 | 0.089 | 0.146 | 0.952 | 11/113/16/19708 |
| BCLVI | 0.999 | 0.333 | 0.053 | 0.091 | 0.895 | 1/18/2/19827 |
| BCLVS | 0.999 | 0.000 | 0.000 | 0.000 | 0.887 | 0/23/3/19822 |
| DH | 1.000 | 0.000 | 0.000 | 0.000 | 0.827 | 0/9/0/19839 |
| Referrable Glaucoma | 0.989 | 0.556 | 0.092 | 0.158 | 0.938 | 20/197/16/19615 |
| LC | 0.995 | 0.111 | 0.012 | 0.022 | 0.951 | 1/82/8/19757 |
| LD | 0.997 | 0.091 | 0.019 | 0.032 | 0.911 | 1/51/10/19786 |
| NVT | 0.998 | 0.000 | 0.000 | 0.000 | 0.956 | 0/31/2/19815 |
| RNFLDI | 0.999 | 0.000 | 0.000 | 0.000 | 0.961 | 0/15/1/19832 |
| RNFLDS | 0.999 | 0.250 | 0.067 | 0.105 | 0.863 | 1/14/3/19830 |

**Table S6d.**
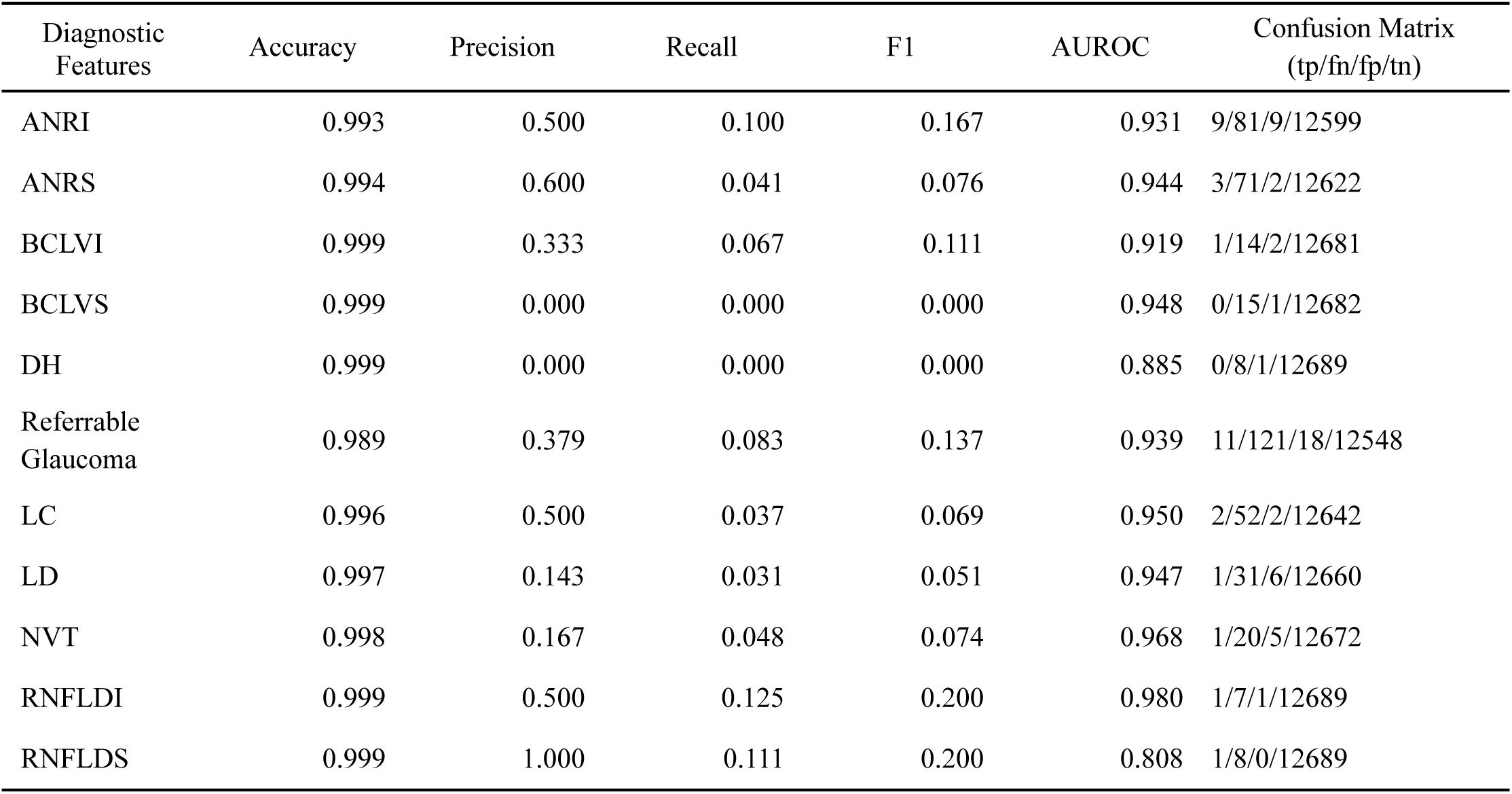
Diagnostic labels classification results for models trained with AIROGS diagnostic features for DINOv2 model trained on AIROGS features and VCDR.

**Table S6e.** Diagnostic labels classification results for models trained with AIROGS diagnostic features for RETFound model trained on AIROGS features.

| Diagnostic Features | Accuracy | Precision | Recall | F1 | AUROC | Confusion Matrix (tp/fn/fp/tn) |
| --- | --- | --- | --- | --- | --- | --- |
| ANRI | 0.993 | 0.643 | 0.061 | 0.112 | 0.923 | 9/138/5/19696 |
| ANRS | 0.994 | 0.600 | 0.097 | 0.167 | 0.934 | 12/112/8/19716 |
| BCLVI | 0.999 | 0.667 | 0.105 | 0.182 | 0.768 | 2/17/1/19828 |
| BCLVS | 0.999 | 0.000 | 0.000 | 0.000 | 0.743 | 0/23/1/19824 |
| DH | 1.000 | 0.000 | 0.000 | 0.000 | 0.402 | 0/9/0/19839 |
| Referrable Glaucoma | 0.989 | 0.545 | 0.083 | 0.144 | 0.926 | 18/199/15/19616 |
| LC | 0.996 | 0.667 | 0.048 | 0.090 | 0.849 | 4/79/2/19763 |
| LD | 0.997 | 0.500 | 0.019 | 0.037 | 0.804 | 1/51/1/19795 |
| NVT | 0.998 | 0.000 | 0.000 | 0.000 | 0.731 | 0/31/1/19816 |
| RNFLDI | 0.999 | 0.500 | 0.133 | 0.211 | 0.828 | 2/13/2/19831 |
| RNFLDS | 0.999 | 1.000 | 0.133 | 0.235 | 0.795 | 2/13/0/19833 |

**Table S6f.** Diagnostic labels classification results for models trained with AIROGS diagnostic features for RETFound model trained on AIROGS features and VCDR.

| Diagnostic Features | Accuracy | Precision | Recall | F1 | AUROC | Confusion Matrix (tp/fn/fp/tn) |
| --- | --- | --- | --- | --- | --- | --- |
| ANRI | 0.993 | 0.444 | 0.178 | 0.254 | 0.859 | 16/74/20/12588 |
| ANRS | 0.991 | 0.267 | 0.270 | 0.268 | 0.920 | 20/54/55/12569 |
| BCLVI | 0.999 | 0.000 | 0.000 | 0.000 | 0.534 | 0/15/2/12681 |
| BCLVS | 0.999 | 0.000 | 0.000 | 0.000 | 0.540 | 0/15/0/12683 |
| DH | 0.999 | 0.000 | 0.000 | 0.000 | 0.465 | 0/8/0/12690 |
| Referrable Glaucoma | 0.988 | 0.402 | 0.265 | 0.320 | 0.905 | 35/97/52/12514 |
| LC | 0.996 | 0.588 | 0.185 | 0.282 | 0.934 | 10/44/7/12637 |
| LD | 0.996 | 0.130 | 0.094 | 0.109 | 0.880 | 3/29/20/12646 |
| NVT | 0.998 | 0.000 | 0.000 | 0.000 | 0.603 | 0/21/0/12677 |
| RNFLDI | 0.999 | 0.000 | 0.000 | 0.000 | 0.665 | 0/8/1/12689 |
| RNFLDS | 0.999 | 0.200 | 0.111 | 0.143 | 0.804 | 1/8/4/12685 |

## Notes

### Competing Interest Statement

The authors have declared no competing interest.

### Funding Statement

This research was supported by the Saudi Data and Artificial Intelligence Authority (SDAIA).

### Summary of Updates

The manuscript has been revised to add Dr. Adi Mohammed Al Owaifeer, as a co-author due to his contributions to the data collection and processing work.

## REFERENCES

Bajwa, Muhammad Naseer, Gur Amrit Pal Singh, Wolfgang Neumeier, Muhammad Imran Malik, Andreas Dengel, and Sheraz Ahmed. 2020. “G1020: A Benchmark Retinal Fundus Image Dataset for Computer-Aided Glaucoma Detection.” *arXiv [eess.IV]*. 10.48550/ARXIV.2006.09158.

Bear Don’t Walk, Oliver J., Iv, Tony Sun, Adler Perotte, and Noémie Elhadad. 2021. “Clinically Relevant Pretraining Is All You Need.” Journal of the American Medical Informatics Association : JAMIA 28 (9): 1970–76.

Burton, Matthew J., Jacqueline Ramke, Ana Patricia Marques, Rupert R. A. Bourne, Nathan Congdon, Iain Jones, Brandon A. M. Ah Tong, et al. 2021. “The Lancet Global Health Commission on Global Eye Health: Vision beyond 2020.” The Lancet. Global Health 9 (4): e489–551.

Chen, Zhihong, Guanbin Li, and Xiang Wan. 2022. “Align, Reason and Learn: Enhancing Medical Vision-and-Language Pre-Training with Knowledge.” In Proceedings of the 30th ACM International Conference on Multimedia. New York, NY, USA: ACM. 10.1145/3503161.3547948.

Diaz-Pinto, Andres, Sandra Morales, Valery Naranjo, Thomas Köhler, Jose M. Mossi, and Amparo Navea. 2019. “CNNs for Automatic Glaucoma Assessment Using Fundus Images: An Extensive Validation.” Biomedical Engineering Online 18 (1): 29.

Gao, Xiaoyi Raymond, Fengze Wu, Phillip T. Yuhas, Rafiul Karim Rasel, and Marion Chiariglione. 2024. “Automated Vertical Cup-to-Disc Ratio Determination from Fundus Images for Glaucoma Detection.” Scientific Reports 14 (1): 4494.

He, Kaiming, Xiangyu Zhang, Shaoqing Ren, and Jian Sun. 2015. “Deep Residual Learning for Image Recognition.” arXiv [cs.CV]. 10.48550/ARXIV.1512.03385.

Hemelings, Ruben, Bart Elen, Alexander K. Schuster, Matthew B. Blaschko, João Barbosa-Breda, Pekko Hujanen, Annika Junglas, et al. 2023. “A Generalizable Deep Learning Regression Model for Automated Glaucoma Screening from Fundus Images.” NPJ Digital Medicine 6 (1): 112.

Kauffmann, Jacob, Jonas Dippel, Lukas Ruff, Wojciech Samek, Klaus-Robert Müller, and Grégoire Montavon. 2025. “Explainable AI Reveals Clever Hans Effects in Unsupervised Learning Models.” Nature Machine Intelligence, March, 1–11.

Kiefer, Riley, Muhammad Abid, Jessica Steen, Mahsa Raeisi Ardali, and Ehsan Amjadian. 2023. “A Catalog of Public Glaucoma Datasets for Machine Learning Applications.” In 2023 the 7th International Conference on Information System and Data Mining (ICISDM), 24–31. New York, NY, USA: ACM.

Kovalyk, Oleksandr, Juan Morales-Sánchez, Rafael Verdú-Monedero, Inmaculada Sellés-Navarro, Ana Palazón-Cabanes, and José-Luis Sancho-Gómez. 2022. “PAPILA.” figshare. 10.6084/M9.FIGSHARE.14798004.V1.

Ling, Xiao Chun, Henry Shen-Lih Chen, Po-Han Yeh, Yu-Chun Cheng, Chu-Yen Huang, Su-Chin Shen, and Yung-Sung Lee. 2025. “Deep Learning in Glaucoma Detection and Progression Prediction: A Systematic Review and Meta-Analysis.” Biomedicines 13 (2). 10.3390/biomedicines13020420.

Liu, Hanruo, Liu Li, I. Michael Wormstone, Chunyan Qiao, Chun Zhang, Ping Liu, Shuning Li, et al. 2019. “Development and Validation of a Deep Learning System to Detect Glaucomatous Optic Neuropathy Using Fundus Photographs.” JAMA Ophthalmology 137 (12): 1353–60.

Li, Zhongwen, Lei Wang, Xuefang Wu, Jiewei Jiang, Wei Qiang, He Xie, Hongjian Zhou, Shanjun Wu, Yi Shao, and Wei Chen. 2023. “Artificial Intelligence in Ophthalmology: The Path to the Real-World Clinic.” *Cell Reports*. Medicine 4 (7): 101095.

Moor, Michael, Oishi Banerjee, Zahra Shakeri Hossein Abad, Harlan M. Krumholz, Jure Leskovec, Eric J. Topol, and Pranav Rajpurkar. 2023. “Foundation Models for Generalist Medical Artificial Intelligence.” Nature 616 (7956): 259–65.

Oquab, Maxime, Timothée Darcet, Théo Moutakanni, Huy Vo, Marc Szafraniec, Vasil Khalidov, Pierre Fernandez, et al. 2023. “DINOv2: Learning Robust Visual Features without Supervision.” arXiv [cs.CV]. 10.48550/ARXIV.2304.07193.

Orlando, José Ignacio, Huazhu Fu, João Barbosa Breda, Karel van Keer, Deepti R. Bathula, Andrés Diaz-Pinto, Ruogu Fang, et al. 2020. “REFUGE Challenge: A Unified Framework for Evaluating Automated Methods for Glaucoma Assessment from Fundus Photographs.” Medical Image Analysis 59 (January):101570.

Schäfer, Raphael, Till Nicke, Henning Höfener, Annkristin Lange, Dorit Merhof, Friedrich Feuerhake, Volkmar Schulz, Johannes Lotz, and Fabian Kiessling. 2024. “Overcoming Data Scarcity in Biomedical Imaging with a Foundational Multi-Task Model.” Nature Computational Science 4 (7): 495–509.

Sivaswamy, Jayanthi, S. R. Krishnadas, Gopal Datt Joshi, Madhulika Jain, and A. Ujjwaft Syed Tabish. 2014. “Drishti-GS: Retinal Image Dataset for Optic Nerve head(ONH) Segmentation.” In 2014 IEEE 11th International Symposium on Biomedical Imaging (ISBI). IEEE. 10.1109/isbi.2014.6867807.

Tan, Rose, Kelvin Yi Chong Teo, Rahat Husain, Ngiap Chuan Tan, Qian Xin Lee, Haslina Hamzah, Tina Wong, et al. 2024. “Evaluating the Outcome of Screening for Glaucoma Using Colour Fundus Photography-Based Referral Criteria in a Teleophthalmology Screening Programme for Diabetic Retinopathy.” The British Journal of Ophthalmology 108 (7): 933–39.

Vente, Coen de, Koenraad A. Vermeer, Nicolas Jaccard, He Wang, Hongyi Sun, Firas Khader, Daniel Truhn, et al. 2024. “AIROGS: Artificial Intelligence for Robust Glaucoma Screening Challenge.” IEEE Transactions on Medical Imaging 43 (1): 542–57.

Xie, Enze, Wenhai Wang, Zhiding Yu, Anima Anandkumar, Jose M. Alvarez, and Ping Luo. 2021. “SegFormer: Simple and Efficient Design for Semantic Segmentation with Transformers.” arXiv [cs.CV]. 10.48550/ARXIV.2105.15203.

Yang Wena, Leiting Chena, b, Yu Dengc, Chuan Zhoua. 2021. “Rethinking Pre-Training on Medical Imaging.” Journal of Visual Communication and Image Representation 78 (July):103145.

Zhang, Chuyan, Hao Zheng, and Yun Gu. 2023. “Dive into the Details of Self-Supervised Learning for Medical Image Analysis.” Medical Image Analysis 89 (October):102879.

Zhang, Zhuo, Feng Shou Yin, Jiang Liu, Wing Kee Wong, Ngan Meng Tan, Beng Hai Lee, Jun Cheng, and Tien Yin Wong. 2010. “ORIGA(-Light): An Online Retinal Fundus Image Database for Glaucoma Analysis and Research.” Annual International Conference of the IEEE Engineering in Medicine and Biology Society. IEEE Engineering in Medicine and Biology Society. Annual International Conference 2010:3065–68.

Zhou, Shihao, Mengxi Jiang, Shanshan Cai, and Yunqi Lei. 2021. “DC-GNet: Deep Mesh Relation Capturing Graph Convolution Network for 3D Human Shape Reconstruction.” arXiv [cs.CV]. 10.48550/ARXIV.2108.12384.

Zhou, Yukun, Mark A. Chia, Siegfried K. Wagner, Murat S. Ayhan, Dominic J. Williamson, Robbert R. Struyven, Timing Liu, et al. 2023. “A Foundation Model for Generalizable Disease Detection from Retinal Images.” Nature 622 (7981): 156–63.

